# The Impact of Climate-Related Extreme Weather on U.S. Health Care Utilization and Costs

**DOI:** 10.1101/2025.06.10.25329348

**Authors:** Shakira J. Grant

## Abstract

**Background:** As global temperatures rise due to the climate crisis, extreme weather events like heatwaves, wildfires, hurricanes, and flooding are becoming more frequent, intense, and deadly. These events increase health risks, including cardiovascular, respiratory, and mental health disorders, which escalate health care utilization and strain the already overburdened United States (U.S.) health care system. Unfortunately, climate denialism does not stop the impact on the U.S. health care facilities or patients. Addressing these issues will improve health and lower health system and patient costs. This rapid evidence review examines U.S.-based studies published between 2018 and 2023 to understand the impact of climate change on health care utilization and costs, identify areas for further research, and discuss policy implications.

**Methods:** A PubMed search identified U.S.-based studies published between 01/01/2018 and 10/31/2023 on health care utilization and costs related to extreme heat, wildfires, and hurricanes/floods. Three independent reviewers extracted key characteristics, including study design, sample size, location, participant sociodemographics, outcomes, and implications for health equity.

**Results:** Out of 554 identified studies, 54 met the inclusion criteria: 21 on heat, 12 on wildfires, and 21 on hurricanes/floods. Geographic patterns emerged, with 38% of extreme heat-related studies focusing on events in the Northeast, 75% of wildfire studies examining trends in the West, and 52% of hurricane/flooding studies using data from the South. Most studies (71%) reported demographics such as age, sex, and race/ethnicity, but none included data on persons with disabilities. One study focused on veterans’ health care utilization before and after Hurricane Harvey. Across all studies, participant ages varied: 33% included all ages, 9% focused on individuals 25 or younger, 11% on adults 18 or older, and 26% on adults 65 and older, with 20% not reporting age. The studies showed that extreme weather events led to increased health care utilization, including emergency department (ED) visits, Emergency Medical Services (EMS) calls, hospitalizations, and outpatient services. Older adults, Black individuals, and those of lower socioeconomic status experienced higher health care utilization. Only two studies examined health care costs, each within one year of Hurricane Sandy (2012) and the 2016 Baton Rouge flooding, both demonstrating increased health care costs during these periods.

**Conclusion:** The evidence is undeniable: climate events– no matter where they occur or whom they impact — drive up health care spending and utilization, straining systems already under pressure. Yet, critical gaps remain. We lack a complete understanding of the long-term consequences, the disproportionate burden on vulnerable populations, and the specific risks faced by individuals with chronic conditions such as cancer, HIV, and kidney disease. The absence of robust data on both immediate and sustained health care costs further weakens our ability to respond effectively. Urgent, targeted research is imperative to fill these gaps, shape strategic interventions, and drive policies that ensure health care systems can withstand and adapt to the escalating challenges climate change poses.

## Introduction

The escalating climate crisis is unleashing record-breaking global temperatures and more frequent, intense extreme weather events, posing profound and growing threats to human health and pushing health care systems to their limits. On July 22, 2024, Earth recorded its hottest day, continuing a trend of rising global temperatures that began in the 1970s.^2^ In the U.S., temperatures have increased by 2.5°F since 1970, outpacing the global rise of 1.7°F.^3^ In 2024, the National Oceanic and Atmospheric Administration (NOAA) reported the hottest year on record, with a global average temperature 2.32°F above the 20th-century average.^3^ As extreme weather events grow more frequent and intense, their health and economic impacts have become an urgent public health crisis.^2,4^

In the U.S., 2024 marked the deadliest, most intense, and costliest tornado season on record. Tornadoes in the central and southern regions between April and May alone caused an estimated$14 billion in damages.^5,6^ More recently, beginning on January 7, 2025, wildfires in the Los Angeles area have devastated communities —destroying infrastructure, displacing thousands, and resulting in at least 28 deaths.^7^

Beyond immediate structural damage, the Los Angeles County Economic Development Corporation Institute for Applied Economics estimated total property losses from these wildfires to range between $28 billion and $53.8 billion.^8^ The same analysis found that many of the affected businesses operated in professional, scientific, and technical services, retail trade, health care, and social assistance.^8^ The projected economic impact includes an estimated $4.6 billion to $8.9 billion in lost economic output between 2025 and 2039, a loss of up to 50,000 job-years, and associated income reductions of $1.9 billion to $ 3.7 billion.^8^ Government tax revenue losses due to employment disruption and reduced business activity are expected to reach $1.5 billion.

Beyond the immediate destruction, climate-driven disasters like wildfires have cumulative health effects. Affected populations often face both short- and long-term health conditions, which carry profound health and economic consequences. These include increased demand for emergency department (ED) visits, hospitalizations, and lost wages.^9^ Given the scale of the California wildfires, the state now stands at a critical juncture—offering a key opportunity to advance research on the short- and long-term health impacts of such events, quantify the financial burden on the health care system, and develop strategies to enhance resilience against future climate-related crises.

Extreme weather events are no longer isolated crises—they are escalating public health emergencies with far-reaching and compounding consequences. Heatwaves pose the greatest climate-driven threat to human health, causing more deaths annually in the U.S. than earthquakes, tornadoes, and floods combined.^10,11^ Heatwave-associated deaths also exceed the number of deaths caused by other non-climate-related events, such as accidental gun incidents, annually.^12^ Between 1979 and 2022, at least 14,000 deaths were directly attributed to extreme heat, a likely underestimate given heat’s role in exacerbating other conditions.^13^ Heat-related deaths among older adults (aged ≥ 65) increased by 85% between the periods 2000–2004 and 2017–2021.^14^ Additionally, wildfires have caused respiratory and cardiovascular health crises and led to mental health disorders such as post-traumatic stress disorder among survivors.^15–18^ Tornadoes have devastated communities, and hurricanes and flooding have increased infectious diseases and worsened chronic health conditions.^15–17,19,20^

Extreme weather events also disrupt health care delivery by damaging infrastructure, causing power outages, and displacing populations.^21^ For example, Hurricane Beryl in 2024 caused severe operational challenges in Houston hospitals, including delayed discharges and overcrowded facilities.^22^ These events not only strain health care systems but also place additional burdens on health care workers, increasing risks of burnout and mental health disorders.^21^ Health care systems must also contend with rising costs: a 2019 study estimated that 10 major climate events resulted in $10 billion in hospital admissions, emergency visits, and lost wages.^23^ Since 1980, climate disasters in the U.S. have cost nearly $2 trillion.^5,24^

Extreme weather events disproportionately impact marginalized populations, as social determinants of health, such as income, race, ethnicity, and disability, limit access to resources and care.^17,25,26^ These groups face higher rates of illness, more severe health complications, and increased mortality during such crises, further deepening existing health inequities.^17,25,26^ Furthermore, the U.S. health care system itself contributes to the climate crisis, accounting for an estimated 8.5% of the nation’s greenhouse gas emissions.^27–29^ These emissions are categorized into three scopes: Scope 1 emissions, which are direct emissions from health care facilities, represent 7% of the total; Scope 2 emissions, which are indirect emissions from the generation of purchased energy, make up 11%; and Scope 3 emissions, which include supply-chain emissions related to medical supplies, devices, and pharmaceuticals, comprise 82% of the total.^29^ These emissions worsen environmental and human health, creating a vicious cycle of increasing health care demand and climate-related harm that results in nearly 100,000 American lives lost annually.^30^

As the climate crisis escalates, understanding health care utilization during and after extreme weather events is essential to mitigate health and economic burdens on individuals, communities, and the health care system. For example, a recent analysis of data from 809,636 Medicare beneficiaries (2008-2019) found that older adults (aged ≥65) exposed to extreme heat, defined as temperatures above the 95th percentile had a 64% higher adjusted risk of heat-related ED visits (HR [95% CI], 1.64 [1.46, 1.85]) and a 4% higher risk of all-cause acute hospitalization (1.04 [1.01, 1.06]) compared to those exposed to temperatures below the 25th percentile.^31^ For individuals under 65, the risk of heat-related ED visits was also higher (2.69 [2.23, 3.23]), as well as the risk of all-cause ED visits (1.03 [1.01, 1.05]).^31^ Nationwide, extreme heat-related ED visits cost $177.3 million per summer, and hospital admissions cost $834.9 million, highlighting the need for targeted interventions to reduce avoidable visits and associated costs.^32^

This paper presents a rapid review and summary of studies published between 2018 and 2023 on the impact of climate-related events on U.S. health care systems. It synthesizes current knowledge, highlights research gaps, evaluates key federal actions, and recommends policy changes to reduce health and economic burdens and address inequities. Although the current political climate has limited funding and reversed many of the previous administration’s climate programs, discouraging some researchers from pursuing work in this area, the urgency of the climate crisis demands continued research. Independent of federal leadership, such research is essential to inform patient care, improve health outcomes, and strengthen the U.S. health care system.

## Methods

### Search Strategy

A medical librarian (A.C.) searched PubMed for articles published between January 1, 2018, and November 2, 2023, using controlled vocabulary and keywords with synonyms that reflected health utilization and cost impacts of extreme heat, flooding, and wildfires. The search included combinations of the following keywords and subject headings: “extreme heat,” “excessive heat,” “heatwave/heat wave,” “heat event,” “heat stress,” “extreme temperature,” “ high temperature,” “hot temperature,” “flooding/floods,” “ hurricane,” “cyclonic storms,” “ tropical storm,” “wildfire, “fire and smoke,” “costs,” “economic,” “utilization,” “expenditure,” “spending,” “emergency department,” “hospitalization,” “admissions,” “inpatient, outpatient,” “Medicare,” “Medicaid.”

### Study Eligibility Criteria and Selection

We included studies that met the following criteria: peer-reviewed scientific articles reporting human research findings related to frequently described extreme weather events, such as extreme heat, wildfires, hurricanes/floods. Additionally, all eligible studies were required to report on the impacts of these events, specifically concerning health care utilization or health care costs. To characterize the U.S. geographical region where each study was conducted, we used the U.S. Census Bureau’s Regions and Divisions State data.^33^

## Results

### Study Characteristics

The PubMed search yielded 554 articles. After applying our eligibility criteria, we excluded studies that did not report on health care utilization or costs.^34–38^ As well as non-original research, including three systematic reviews/meta-analyses and two narrative or scoping reviews. This resulted in a final analytic sample of 54 articles included in this review.

#### Study Design by Extreme Weather Event

Of these 54 articles, 21 (39%) focused on extreme heat,^39–59^ 12 (22%) on wildfires,^60–71^ and 21 (39%) on hurricanes/floods.^72–92^ Most of these studies were published in 2021,^44,46,47,49,54,57,58,65–67,75,77,78,80–84^ as shown in **Figure 1**. For studies reporting on hurricanes, Sandy ^93^ (a 2012 hurricane that impacted the Eastern Mid-Atlantic region) and Harvey (a 2017 hurricane that affected the Southeastern U.S.) were the most studied hurricanes in seven^73,76,80,84,85,89,92^ and six studies,^72,75,77,81,86,87^ respectively, as shown in **Figure 2**.

**Figure 1:**
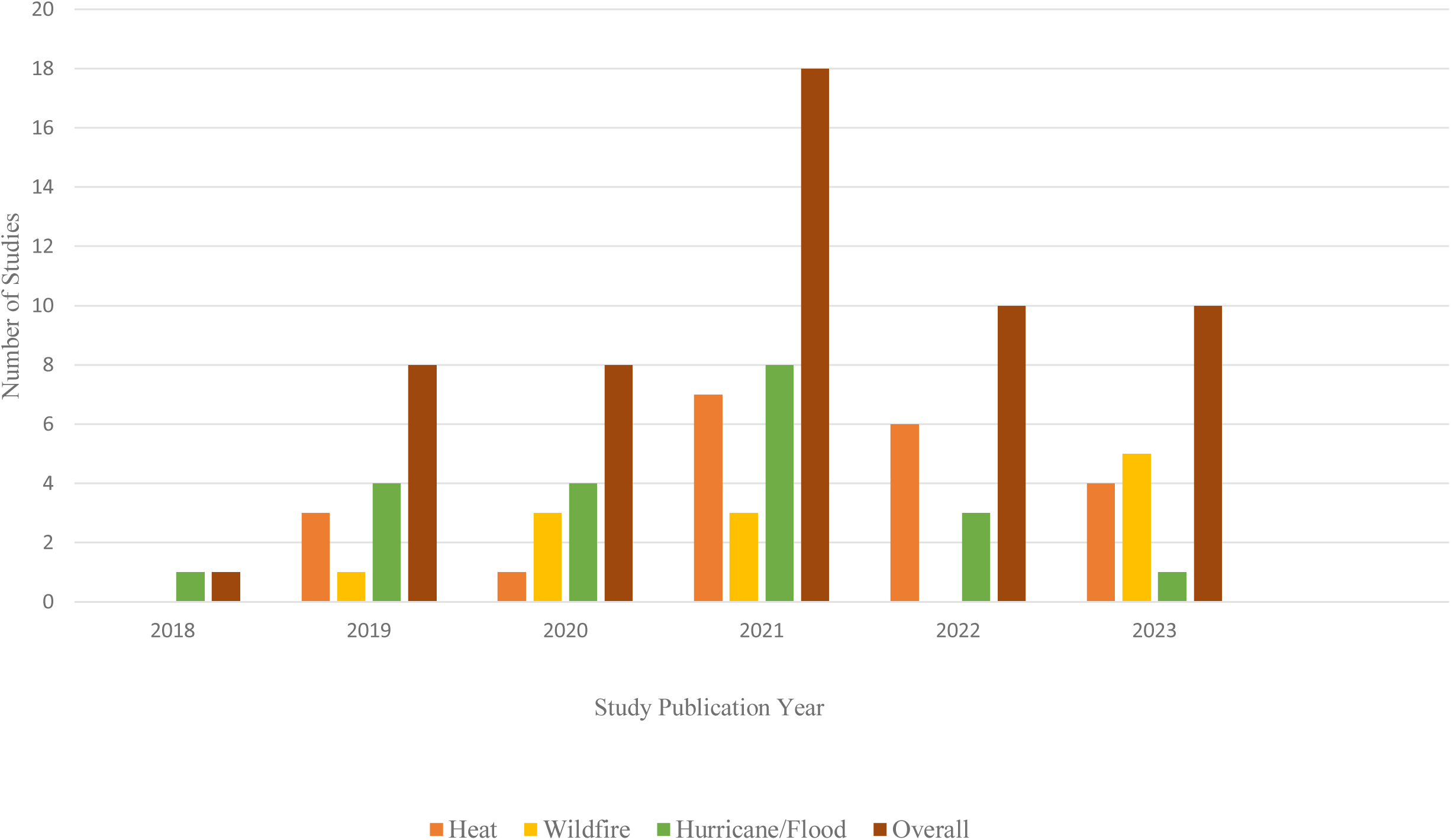
Study Publication Year Acccording to Climate-Related Event. Footnote: illustrates the number of studies published between 2018 and 2023, categorized by climate-related events—heat, wildfires, and hurricanes/floods. The overall number of studies on climate-related health impacts increased over time, with a notable peak in 2021. Studies focusing on extreme heat and wildfires saw a significant rise in 2021 and 2022, while research on hurricanes and floods remained relatively stable.

**Figure 2.**
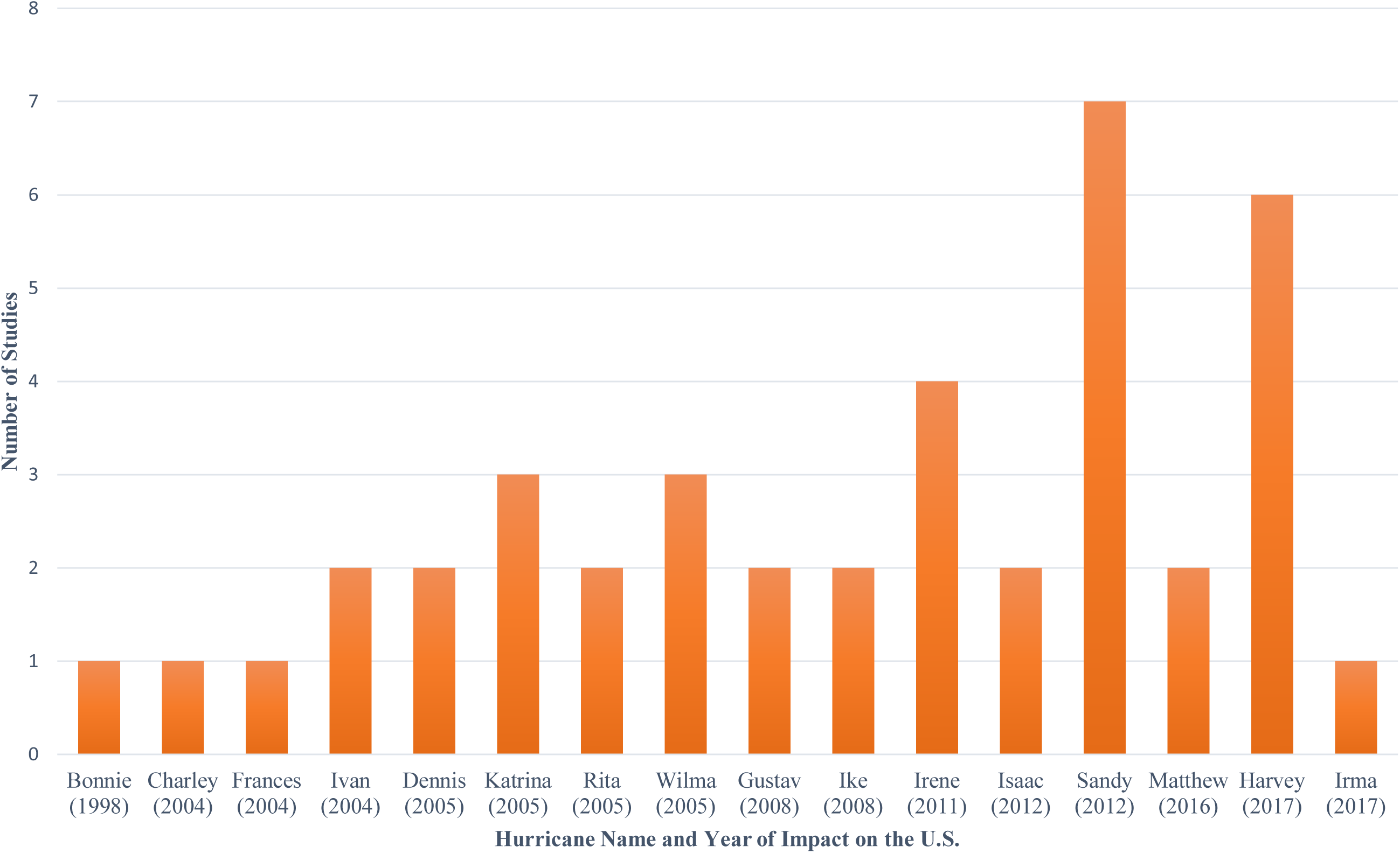
Frequency of Hurricanes Studied. Footnote: displays the frequency of studies examining various U.S. hurricanes from 1998 to 2017. The data show that hurricanes with widespread devastation, such as Hurricane Sandy (2012) and Hurricane Harvey (2017), have been the most frequently studied, followed by Hurricane Irene (2011) and Hurricane Katrina (2005). In contrast, earlier and less catastrophic hurricanes, such as Bonnie (1998) and Charley (2004), have been the focus of fewer studies.

Figure 3 presents the study designs, categorized by the type of climate-related event across all 54 studies. The included studies consisted of 16 case-crossover studies,^41,43–45,51,53–56,58,63–66,70^ 17 retrospective cohort studies,^50,52,59–61,69,72,74,76,79,81,85,86,88–91^ six time-series analyses,^39,40,57,62,67,68^ and two case-control studies.^82,83^ Additionally, five studies employed different methods: case series,^84^ survey,^73^ cross-sectional,^77^ mixed-methods,^87^ or pre-post-test designs.^75^ The study design was not reported for eight studies (four focused on heat-related events,^42,46,47,49^ one focused on wildfires,^71^ and three focused on hurricanes/floods^78,80,92^).

**Figure 3.**
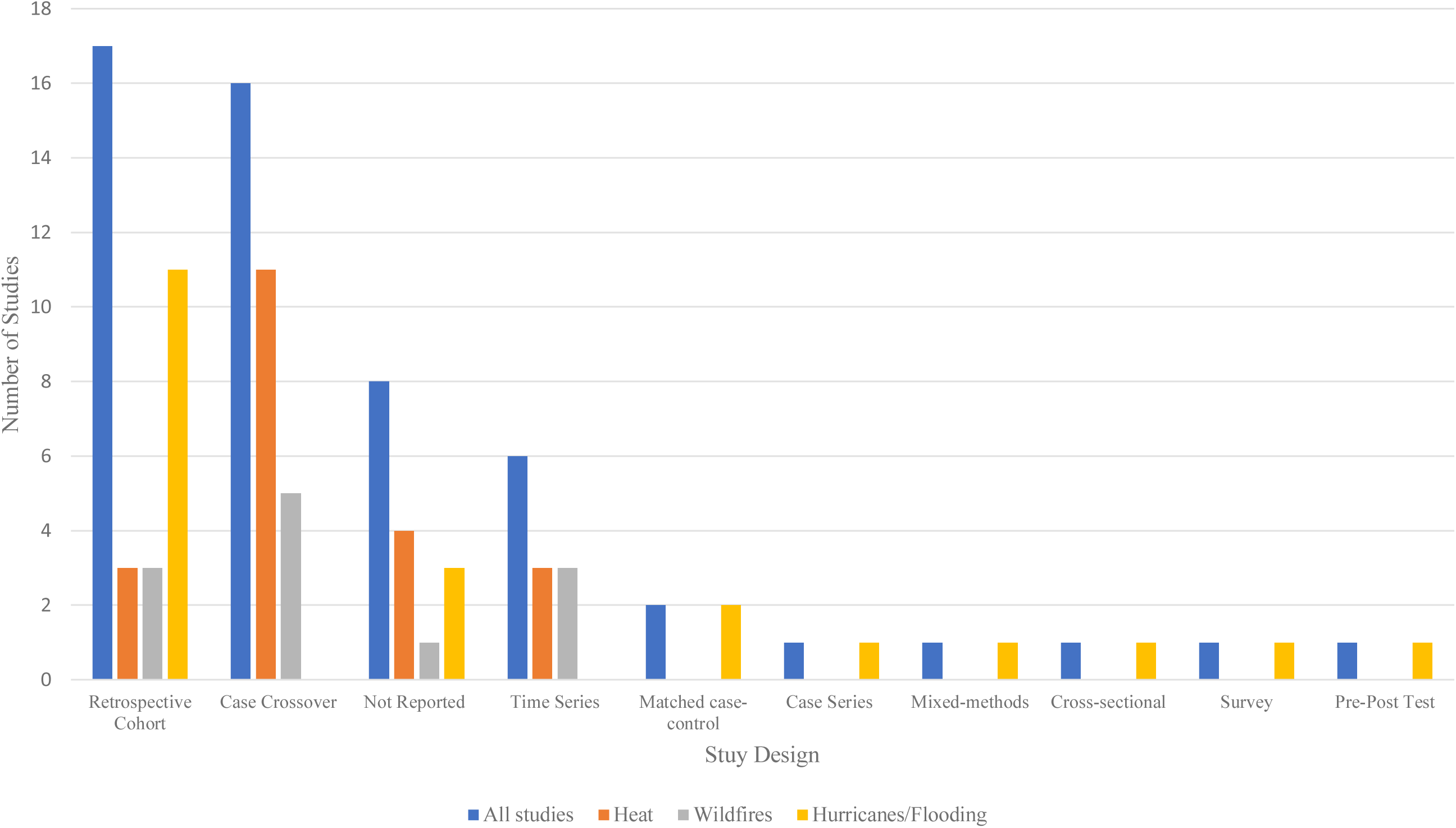
Study Designs of N=54 studies. Footnote: presents the distribution of study designs among the 54 studies reviewed, categorized by climate-related events (heat, wildfires, and hurricanes/floods). Retrospective cohort and case-crossover designs were the most commonly used methodologies. A significant number of studies did not report their study design, highlighting a potential gap in methodological transparency. Time-series analyses were also frequently employed, particularly for evaluating trends over time. Less commonly used designs, such as mixed-methods, cross-sectional studies, surveys, and pre-post tests, suggest that qualitative and observational approaches are underutilized in climate-related health research.

#### Geographic Pattern of Extreme Weather Events

Climate-related events in the U.S. followed a distinct geographic pattern. For extreme heat, eight studies (38%) were conducted in the Northeastern U.S.,^43,46,47,52,53,55–57^ and seven studies (33%) were conducted across multiple U.S. regions,^39–41,48,49,58,59^ as shown in **Table 1**. For wildfires, 9 out of 12 studies (75%) were conducted in the Western U.S. alone.^61–65,67–70^ Meanwhile, 11 of 21 studies (52%) reporting on hurricanes or flooding were undertaken in the Southern U.S.^72,75,77–79,81,86–88,90,91^

**Table 1.** Study Characteristics.

| **Study or Participant Characteristic** | **Heat** | **Wildfires** | **Hurricanes** | **Total** |
| --- | --- | --- | --- | --- |
| Final number of original articles included | 21 | 12 | 21 | 54 |
| **Demographics** |  |  |  |  |
| All ages | 3 | 7 | 8 | 18 |
| 0-25 | 4 | 0 | 1 | 5 |
| 18+ | 4 | 1 | 1 | 6 |
| 65+ | 5 | 1 | 8 | 14 |
| Age not reported- deidentified data | 5 | 3 | 3 | 11 |
| **Total** | 21 | 12 | 21 | 54 |
| **Publication Year** |  |  |  | 0 |
| 2018 | 0 | 0 | 1 | 1 |
| 2019 | 3 | 1 | 4 | 8 |
| 2020 | 1 | 3 | 4 | 8 |
| 2021 | 7 | 3 | 8 | 18 |
| 2022 | 6 | 0 | 3 | 9 |
| 2023 | 4 | 5 | 1 | 10 |
| **Total** | 21 | 12 | 21 | 54 |
| **Study Design** |  |  |  |  |
| Case crossover | 11 | 5 | 0 | 16 |
| Case series | 0 | 0 | 1 | 1 |
| Cross-sectional | 0 | 0 | 1 | 1 |
| Matched case-control | 0 | 0 | 2 | 2 |
| Mixed-methods | 0 | 0 | 1 | 1 |
| Pre-post test | 0 | 0 | 1 | 1 |
| Retrospective cohort | 3 | 3 | 11 | 17 |
| Survey | 0 | 0 | 1 | 1 |
| Time series | 3 | 3 | 0 | 6 |
| Not reported | 4 | 1 | 3 | 8 |
| **Total** | 21 | 12 | 21 | 54 |
| **Study Population** |  |  |  |  |
| Medicare Beneficiaries |  |  |  |  |
| Yes | 8 | 3 | 9 | 20 |
| No | 9 | 9 | 12 | 20 |
| Unable to determine | 4 | 0 | 0 | 4 |
| **Total** | 21 | 12 | 21 | 54 |
| **Study Region** |  |  |  |  |
| United States Census Bureau Regions |  |  |  |  |
| West | 3 | 9 | 0 | 12 |
| Midwest | 0 | 0 | 0 | 0 |
| Northeast | 8 | 1 | 4 | 13 |
| South | 3 | 0 | 11 | 14 |
| Multiple regions | 7 | 2 | 6 | 15 |
| **Total** | 21 | 12 | 21 | 54 |
| **Outcomes Assessed** |  |  |  |  |
| Emergency Department/Urgent<br>Care Center Visits | 11 | 9 | 13 | 33 |
| Emergency Medical Services Calls | 2 | 0 | 0 | 2 |
| Hospitalizations | 8 | 2 | 10 | 20 |
| Outpatient Clinic Visits | 1 | 1 | 0 | 2 |
| Health Care Costs | 0 | 0 | 2 | 2 |

### Study Participant Characteristics

The sample sizes of individual studies and the datasets used to conduct data analyses were highly variable. Some studies leveraged the medical records of thousands of patients, while others examined administrative claims data for millions of beneficiaries. **Table 2** summarizes the datasets reported per individual study.

**Table 2:** List of Datasets Reported by Each Included Study.

| Weather event | First Author | Dataset |
| --- | --- | --- |
| Heat | Aaron S. Bernstein | Pediatric Health Information System (PHIS) |
|  | Alexander Liss | Medicare Provider Analysis and Review (MEDPAR) Files |
|  |  | Centers for Medicare & Medicaid Services set of annual demographic summaries on Medicare beneficiaries |
|  |  | National Oceanic and Atmospheric Administration Global Summary of The Day (NOAA-GSOD) |
|  | Amruta Nori-Sarma | OptumLabs Data Warehouse |
|  | Amruta Nori-Sarma | OptumLabs Data Warehouse |
|  | Blean Girma | New York Statewide Planning and Research Cooperative System |
|  | Daniel Samano | National Oceanic and Atmospheric Administration Global Summary of The Day (NOAA-GSOD) |
|  |  | HIV Clinic Registry Database (Registry housed in a large county hospital in South Florida) |
|  | Eric L. Landguth | Hospital Registry Data (Registry housed at a Missoula County, Montana hospital) |
|  | Eun-Hye Yoo | New York Statewide Planning and Research Cooperative System |
|  |  | National Center for Environmental Information Climate Data Online System |
|  | Eun-Hye Yoo | New York Statewide Planning and Research Cooperative System |
|  |  | National Center for Environmental Information Climate Data Online System |
|  | J Bradley Layton | Centers for Medicare & Medicaid Services Claims Data |
|  | Kate R Weinberger | National Oceanic and Atmospheric Administration (NOAA) 2016 Data |
|  |  | Centers for Medicare & Medicaid Services Claims Data |
|  | Kijin Seong | Austin-Travis County Emergency Services (EMS incident data) |
|  |  | U.S. Climate Reference Network (weather data) |
|  | Lara Schwarz | EPIC Electronic Health Data (Epic, Verona, WI) |
|  |  | Parameter-elevation Relationships on Independent Slopes Model (PRISM) Climate Group of Oregon State University |
|  | Leah Santacroce | Massachusetts General Brigham Research Patient Data Registry |
|  |  | Centers for Disease Control and Prevention/The Agency for Toxic Substances and Disease Registry's Social Vulnerability Index (SVI) |
|  | Li Niu | New York Statewide Planning and Research Cooperative System |
|  |  | National Oceanic and Atmospheric Administration National Climate Data Center (NCDC) |
|  | Lisa K Zottarelli | National Oceanic and Atmospheric Administration Health Index (HI) Data |
|  |  | Centers for Disease Control and Prevention/The Agency for Toxic Substances and Disease Registry's Social Vulnerability Index (SVI) |
|  | Mengxuan Li | New York Statewide Planning and Research Cooperative System |
|  |  | National Centers for Environmental Information (NCEI) |
|  | Penelope Dring | Nationwide Emergency Department Sample from the Agency for Healthcare Research and Quality |
|  |  | National Oceanic and Atmospheric Administration Climate at a Glance |
|  | Richard V Remigio | Fresenius Kidney Care Clinics (hospital admission and mortality records) |
|  | Seulkee Heo | 2010 U.S. Census Bureau Data |
|  |  | Centers for Medicare & Medicaid Services claims data. |
|  |  | Parameter-elevation Relationships on Independent Slopes Model (PRISM) |
|  | Shenghzi Sun | OptumLabs Data Warehouse |
|  |  | Parameter-elevation Relationships on Independent Slopes Model (PRISM) |
| Wildfires | Kai Chen | NYC Syndromic Surveillance System (Asthma-related ED visits) |
|  |  | Environmental Protection Agency (ambient particulate matter data) |
|  | Sam Heft-Neal | California's Department of Health Care Access and Information (HCAI) |
|  |  | American Community Survey |
|  | Annie I Chen | United States Environmental Protection Agency (EPA) Air Quality System Data Mart |
|  |  | NOAA Hazard Mapping System (HMS) |
|  |  | California Department of Forestry and Fire Protection's Fire and Resource Assessment Program |
|  |  | California Irrigation Management Information System (CIMIS) |
|  |  | National Centers for Environmental Information (NCEI) |
|  |  | American Community Survey |
|  |  | California Department of Health Care Access and Information (ED data and patient discharge data) |
|  | Annie Doubleday | Air Indicator Report for Public Awareness and Community Tracking (AIRPACT-4) model domain |
|  |  | National Oceanic and Atmospheric Administration's (NOAA) daily hazard mapping system |
|  |  | Rapid Health Information Network (RHINO) |
|  | Holly Elser | Optum Clinformatics® Data Mart Database with de-identified commercial and MA claims |
|  |  | Environmental Protection Agency Air Quality System (AQS), |
|  |  | Gridded Surface Meteorological dataset (gridMET) |
|  | M B Hahn | Alaska Department of Environmental Conservation (DEC) for Environmental Protection Agency (EPA) regulatory monitoring |
|  |  | National Oceanic and Atmospheric Administration's (NOAA) Automated Surface Observing System (ASOS) |
|  |  | Alaska Health Facilities Data Reporting Program (HFDR) |
|  | Cecilia Sorensen | National-scale proprietary database owned by Premier, Inc. |
|  |  | National Oceanic and Atmospheric Administration's (NOAA) Hazard Mapping System (HMS) |
|  |  | U.S. Environmental Protection Agency Air Quality System (AQS) |
|  |  | National Oceanic and Atmospheric Administration's (NOAA) National Climate Data Center |
|  | Joan A Casey | Shasta County Health and Human Services Agency |
|  |  | National Oceanic and Atmospheric Administration's (NOAA) Hazard Mapping System (HMS) |
|  | Rosana Aguilera | Fire and Resource Assessment Program |
|  |  | Moderate Resolution Imaging Spectroradiometer (MODIS) Rapid Response System |
|  |  | National Oceanic and Atmospheric Administration's (NOAA) Hazard Mapping System (HMS) |
|  |  | Office of Statewide Health Planning and Development database |
|  | Daniel Kiser | Washoe County Health District Air Quality Management Division (WCHD-AQMD) |
|  |  | Renown's Electronic Health Record for two Emergency Departments and seven Urgent Care Centers |
|  | Ryan Gan | U.S. Environmental Protection Agency Air Quality System (AQS) |
|  |  | National Environmental Satellite, Data, and Information Service (NESDIS) Hazard Mapping System (HMS) |
|  |  | Oregon All Payer All Claims Database (APAC) |
|  | Stephanie DeFlorio-Barker | Medicare Provider Analysis and Review (MEDPAR) files from the Center for Medicare and Medicaid Services (CMS) |
| **Hurricanes/floods** | Yiyao Li | Texas Health Care Information Council (THCIC) |
|  |  | Texas Behavioral Risk Factor Surveillance System (BRFSS) Public Use Data File |
|  | Laura Sands | Center for Medicare and Medicaid Services (CMS) Claims Data |
|  | Laura Sands | Ongoing Research on Aging in New Jersey: Bettering Opportunities for Wellness in Life (ORANJ BOWL) panel study database |
|  | Victoria Lynch | National Inpatient Sample (NIS) from the Healthcare Cost and Utilization Project (HCUP) |
|  |  | NASA/ NOAA North American Land Data Assimilation System 2 (NLDAS-2) |
|  |  | United States Geological Survey (USGS) National Water Information System |
|  |  | National Oceanic and Atmospheric Administration's (NOAA) Storm Event Database |
|  | Balaji Ramesh | Texas Department of Health and Human Services data |
|  |  | NASA MODIS, ESA Sentinel 1, ASI Cosmo SkyMed, and Radarsat 2 data |
|  |  | Dartmouth Flood Observatory data |
|  | Kevin Heslin | Healthcare Cost and Utilization Project State Inpatient Databases and State Emergency Department Databases |
|  | S Aya Fanny | Diagnostic medical records from Quaternary Care Children's Hospital in Houston, Texas |
|  | Kristen Cowan | Healthcare Cost and Utilization Project State Emergency Department and Inpatient Databases |
|  | Cassandra Hua | Medicare Master Beneficiary Summary File (MBSF) |
|  |  | Residential History File (RHF) |
|  |  | ZIP Code History file (ZHF) |
|  |  | List of licensed AL communities |
|  |  | Medicare Provider Analysis and Review (MEDPAR) inpatient claims |
|  |  | Centers for Medicare & Medicaid Services Claims Data |
|  | Kate Weinberger | New York Statewide Planning and Research Cooperative System |
|  | Margaret Carrel | Veterans Affairs Corporate Data Warehouse |
|  | Meilin Yan | Centers for Medicare & Medicaid Services Claims Data |
|  | Robbie Parks | Centers for Medicare & Medicaid Services Claims Data |
|  | Sue Anne Bell | Centers for Medicare & Medicaid Services Claims Data |
|  | Audrey Weiss | State Inpatient Databases (SID) |
|  |  | State Emergency Department Databases (SEDD) |
|  | Cina Karimaghaei | Electronic Health Records |
|  | Kimberly Chambers | Electronic Health Records |
|  | Stephen Phillippi | Centers for Medicare & Medicaid Services Claims Data (Medicaid) |
|  | Wayne Lawrence | Office of the Assistant Secretary of Preparedness and Response Enclave |
|  |  | Centers for Medicare & Medicaid Services Claims Data |
|  | Troy Quast | Centers for Medicare & Medicaid Services Enrollment and Encounter Claims Data |
|  | Troy Quast | Centers for Medicare & Medicaid Services Claims Data |
|  | Linda McQuade | Agency for Healthcare Research and Quality (AHRQ) Healthcare Cost and Utilization (HCUP) |

Most (71%) of the studies reported the individual demographics of enrolled study participants. The demographics most frequently reported were age, sex, and race/ethnicity. None of the studies in the review reported demographic data for people with disabilities. One study exclusively focused on veterans, examining health care utilization within the Veterans Health Administration before and after Hurricane Harvey.^81^ The ages of study participants varied across studies. Nineteen (35%) of the studies included individuals of all ages,^46,47,59–63,65,66,70,72,75,76,78,80,85–88^ while five (9%) focused specifically on persons 25 years or younger,^39,43,45,53,77^ six (11%) focused on those aged ≥ 18,^41,51,52,58,64,81^ and 14 (26%) on adults aged 65 and older.^40,48,49,55,57,71,73,79,82–84,90–92^ Age was not reported in 10 (19%) of the studies.^42,44,50,54,56,67–69,74,89^

#### Extreme Weather Events and Medicare Beneficiaries

Twenty studies (39%)^40–42,48,49,55,57,58,64,66,71,73,78,79,82–84,89–91^ enrolled Medicare populations (see Figure 4). In most studies (80%), Medicare beneficiaries were exclusively 65 years and older, ^40,41,48,49,55,57,58,64,71,73,78,79,82–84,89–91^ while in two studies, the age of the beneficiaries could not be determined. Four studies, ^46,47,50,54^ all focused on extreme heat and did not specify whether Medicare beneficiaries were enrolled. Most studies (n=14) ^40,48,49,55,57,66,71,73,78,79,82,83,90,91^ included those enrolled in traditional Medicare. Among these, seven studies^73,78,79,82,83,90,91^ focused on hurricanes/floods, five on extreme heat, ^40,48,49,55,57^ and two on wildfires.^66,71^ One study focused solely on Medicare Advantage (MA) enrollees.^41^ Additionally, two studies enrolled individuals dually eligible for Medicare and Medicaid,^84,89^ and three studies examined populations enrolled in both MA and commercial plans.^42,58,64^

**Figure 4.**
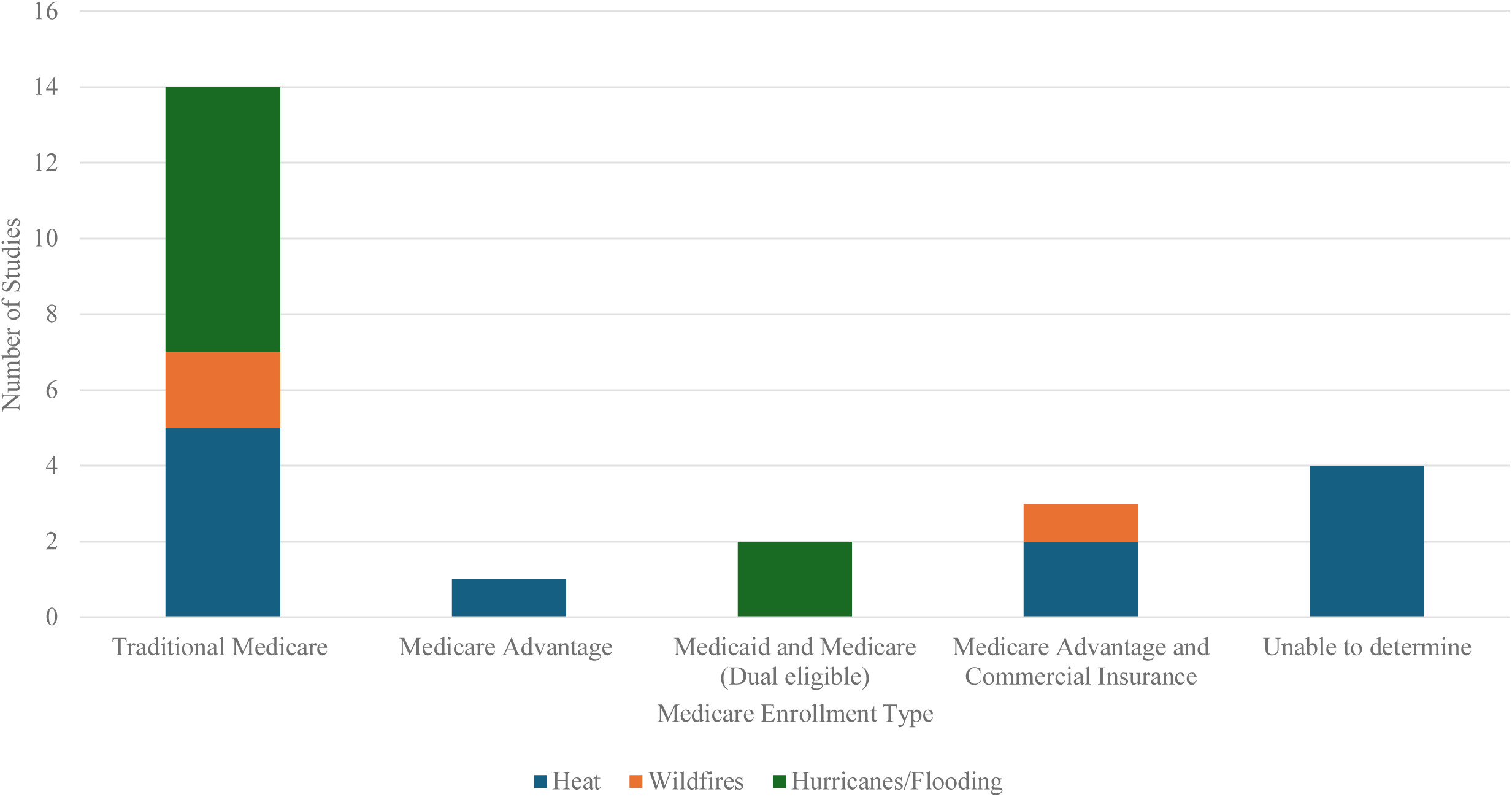
Studies with Enrolled Medicare Beneficiaries. Footnote: presents the distribution of studies that included Medicare beneficiaries, categorized by enrollment type and climate-related event (heat, wildfires, and hurricanes/floods). The majority of studies focused on beneficiaries enrolled in Traditional Medicare, with relatively fewer studies examining those in Medicare Advantage or dual-eligible Medicaid and Medicare populations. Research on Medicare beneficiaries affected by heat-related events was the most prevalent, particularly within Traditional Medicare. A notable proportion of studies did not specify the type of Medicare enrollment, highlighting a gap in data transparency.

### Understanding Extreme Weather Event Impacts on Health Care Utilization and Costs

#### Health Care Utilization: Emergency Services

Extreme weather events led to an increased demand for emergency care services, hospitalization rates, and outpatient primary or specialty provider services, as shown in Figure 5. The definition of emergency care services varied across studies, with two studies including the use of emergency medical services (EMS) call data.^50,54^ Both of these studies focused on heat-related events.^50,54^ The remaining 33 studies^39,41–43,46,47,51,53,55,58–65,67,69,70,75–81,85–87,89,91,92^ focused on visits to either an ED or an Urgent Care Center. Wildfire-related studies examined ED visit utilization more frequently (75%) than hurricane/flood-related studies (62%) and heat-related studies (52%), as shown in Figure 6.

**Figure 5.**
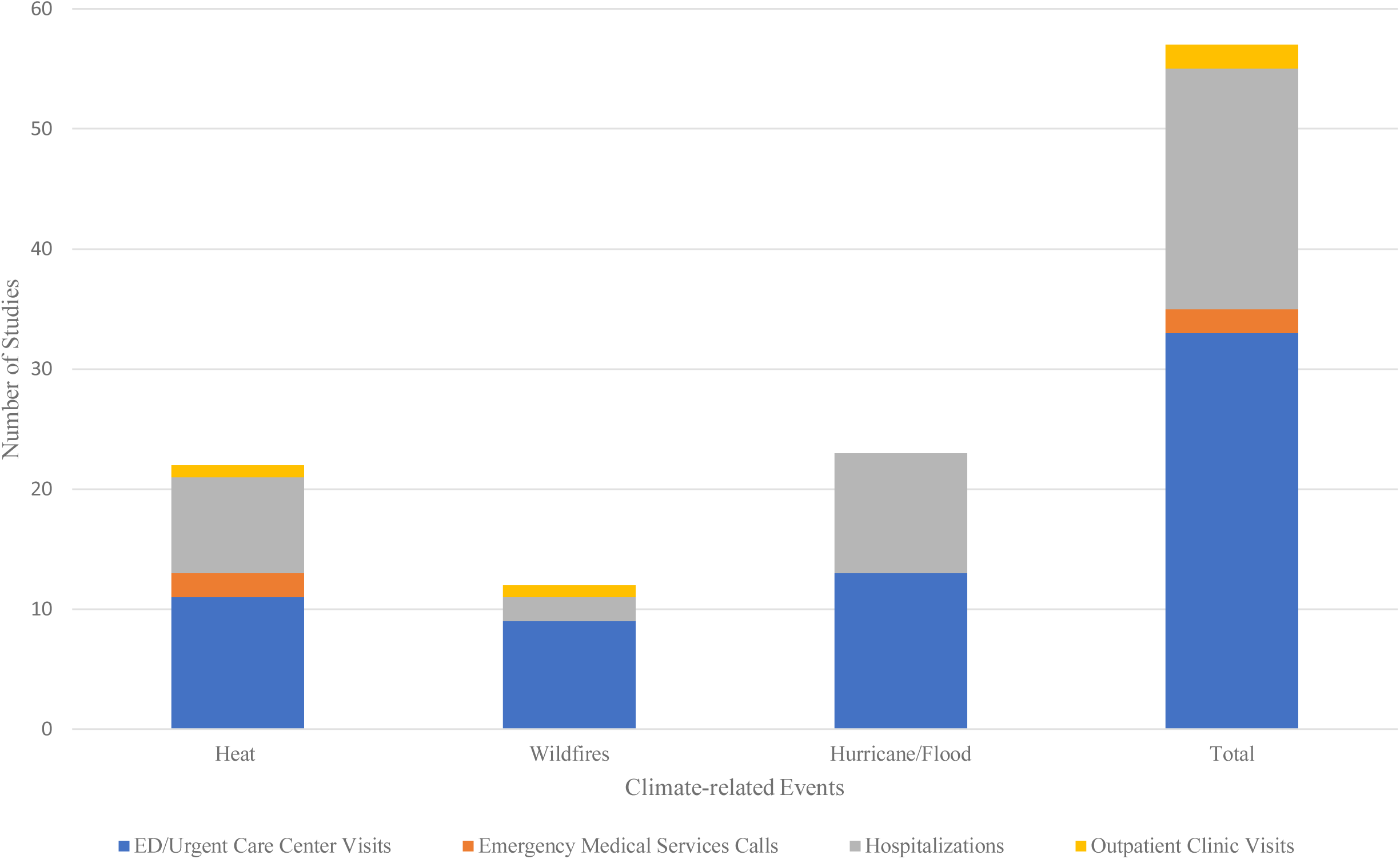
Health Care Utilization According to Climate-Related Events. Footnote: illustrates health care utilization across different climate-related events, including heat, wildfires, and hurricanes/floods. The majority of studies reported increases in emergency department (ED) and urgent care center visits, particularly in response to heat-related events, followed by hurricanes and floods. Hospitalizations also constituted a significant portion of health care utilization, especially for hurricanes and floods, reflecting the severe health impacts of these disasters. Emergency medical services (EMS) calls and outpatient clinic visits were reported less frequently across studies.

**Figure 6.**
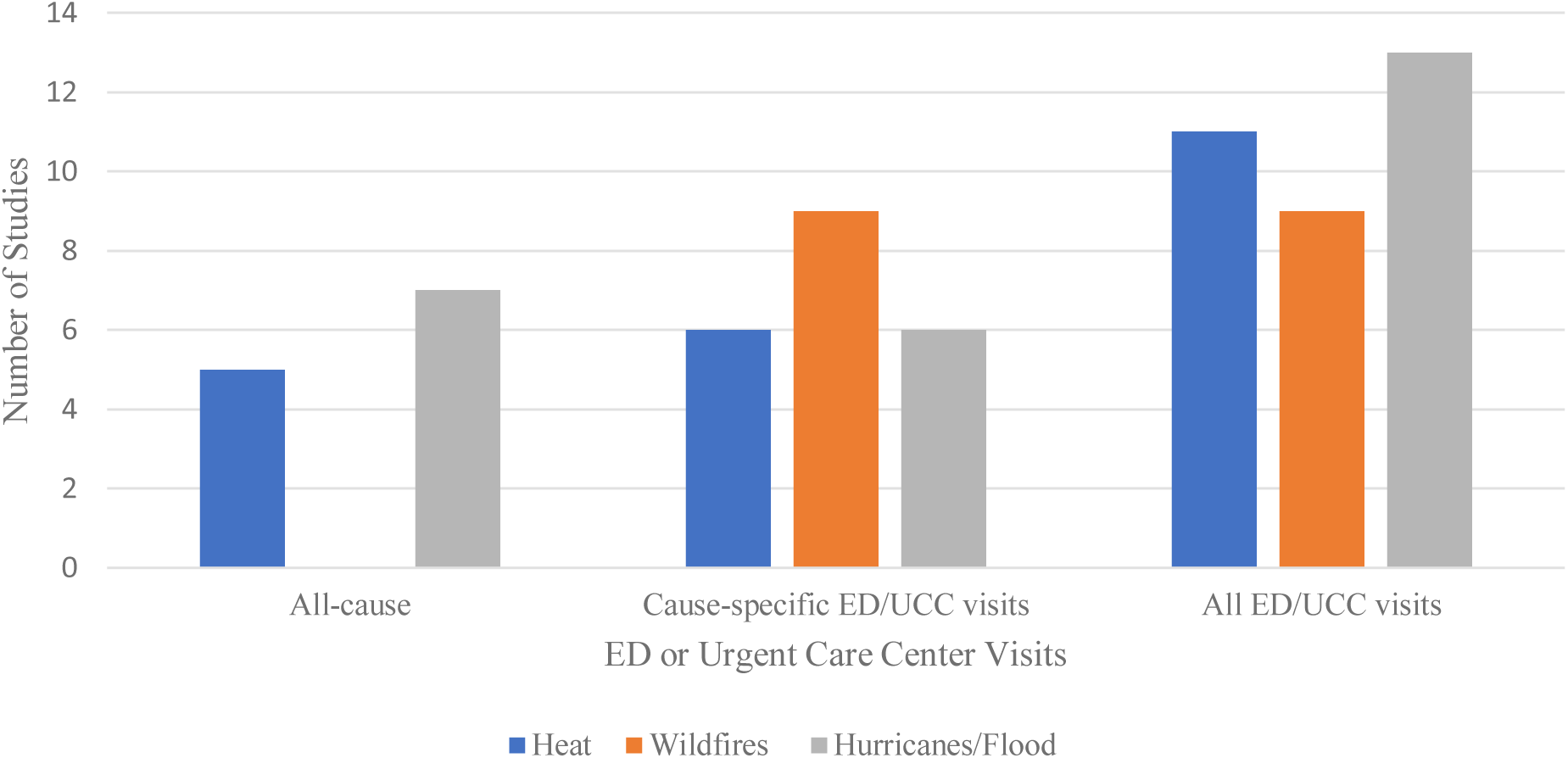
ED or Urgent Care Center Visits by Climate-related Events. Footnote: illustrates the number of studies reporting emergency department (ED) or urgent care center (UCC) visits associated with different climate-related events, categorized by all-cause visits, cause-specific visits, and total ED/UCC visits. Studies on hurricanes and flooding reported the highest number of total ED/UCC visits, followed closely by heat-related events. Cause-specific visits, which focus on particular health conditions triggered by climate events, were most frequently reported in studies on wildfires, likely reflecting the respiratory and cardiovascular impacts of wildfire smoke exposure.

#### Cause-Specific Emergency Services Utilization

Among the 33 studies^39,41–43,46,47,51,53,55,58–65,67,69,70,75–81,85–87,89,91,92^ reporting on ED utilization, 12 examined ED visits related to any cause,^39,42,51,53,58,75–77,79,81,87,91^ while 21 examined cause-specific visits.^41,43,46,47,55,59–65,67,69,70,78,80,85,86,89,92^ Two heat-related studies leveraging EMS call data characterized utilization based on all causes.^50,54^ The reasons for ED visits, categorized by organ systems and as adjudicated by each study, are displayed in Figure 7. Six studies (three each focused on wildfires^62,65,67^ and hurricanes/floods,^80,89,92^) examined multiple cause-related ED visits, most often the presence of both cardiovascular and respiratory-related conditions.

**Figure 7.**
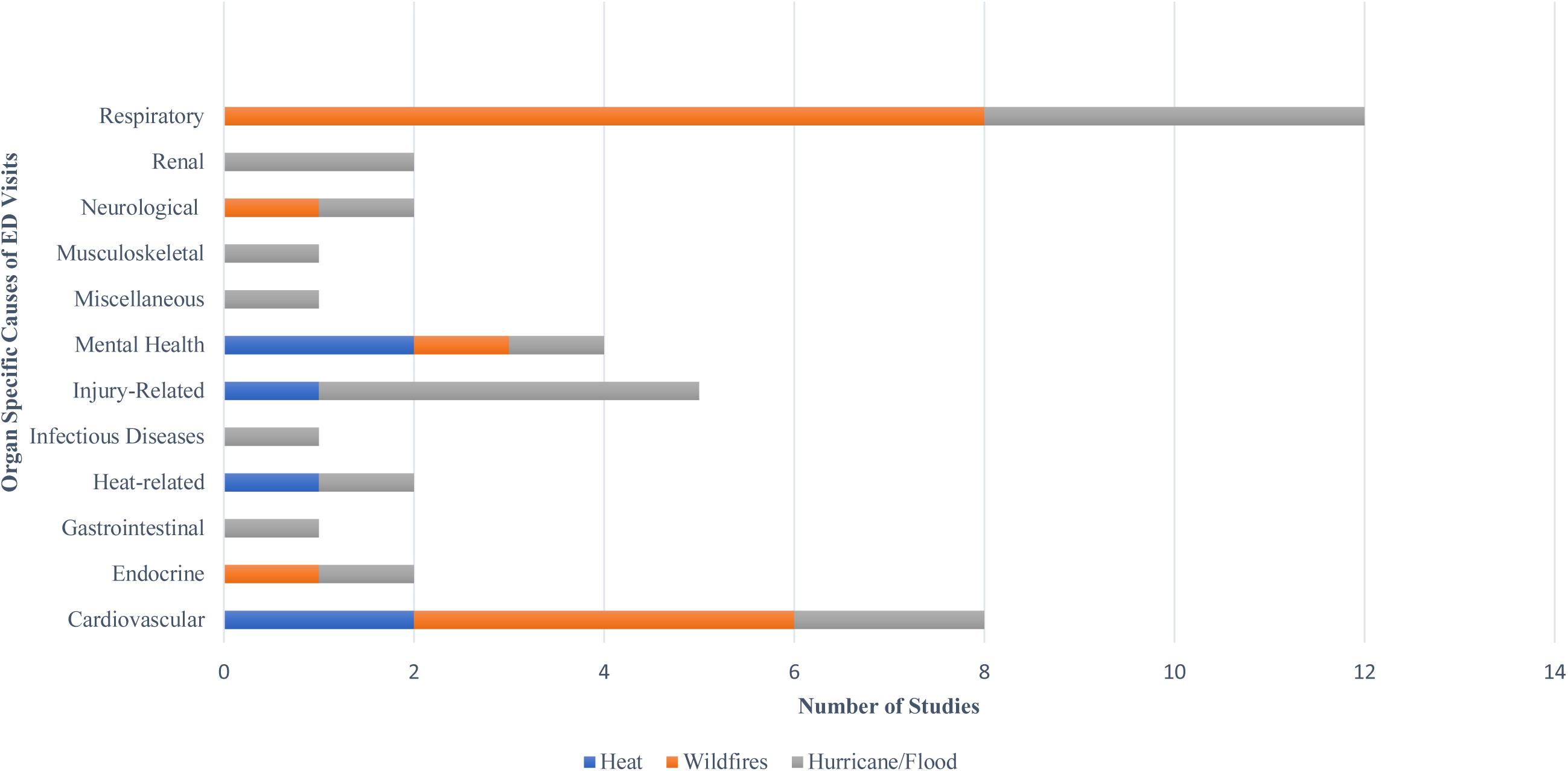
Cause-specific ED Visits. Footnote: presents the distribution of cause-specific emergency department (ED) visits associated with different climate-related events, categorized by organ-specific health impacts. Respiratory conditions were the most frequently studied cause of ED visits, particularly in relation to wildfires and hurricanes/floods, likely due to wildfire smoke exposure and mold-related respiratory issues following flooding events. Cardiovascular conditions were also commonly reported across all climate events, with heat and wildfire exposure contributing significantly. Mental health-related ED visits were associated with all climate events, underscoring the psychological toll of extreme weather disasters. Other notable causes included injury-related visits (frequent in hurricanes and flooding), heat-related illnesses (specific to heat events), and infectious diseases (primarily following hurricanes and floods).

Overall, most of the cause-specific ED visits were due to respiratory conditions, which were frequently studied in the context of wildfires (eight studies),^60–63,65,67,69,70^ or hurricane/flooding (four studies).^78,80,89,92^, The limited reporting of respiratory-related visits during extreme heat events suggests a research gap, and future studies could provide valuable insights by focusing on this area.

#### Health Care Utilization: Hospitalizations

Among the 20 studies^40,45,48,49,52,53,56,57,68,71–74,81–84,89,91,92^ evaluating health care utilization according to acute inpatient hospitalizations, which also included admission to critical care units, shown in Figure 8—most (10 studies)^72–74,81–84,89,91,92^ were related to hurricanes/floods, followed by eight studies reporting on heat-related events,^40,45,48,49,52,53,56,57^ and two on wildfires.^68,92^ Eight studies shown in Figure 8 examined all-cause hospitalizations (defined as hospitalization from any cause); these were related to extreme heat (three studies)^52,53,56^ and hurricanes/floods (five studies).^73,81,82,84,91^

**Figure 8.**
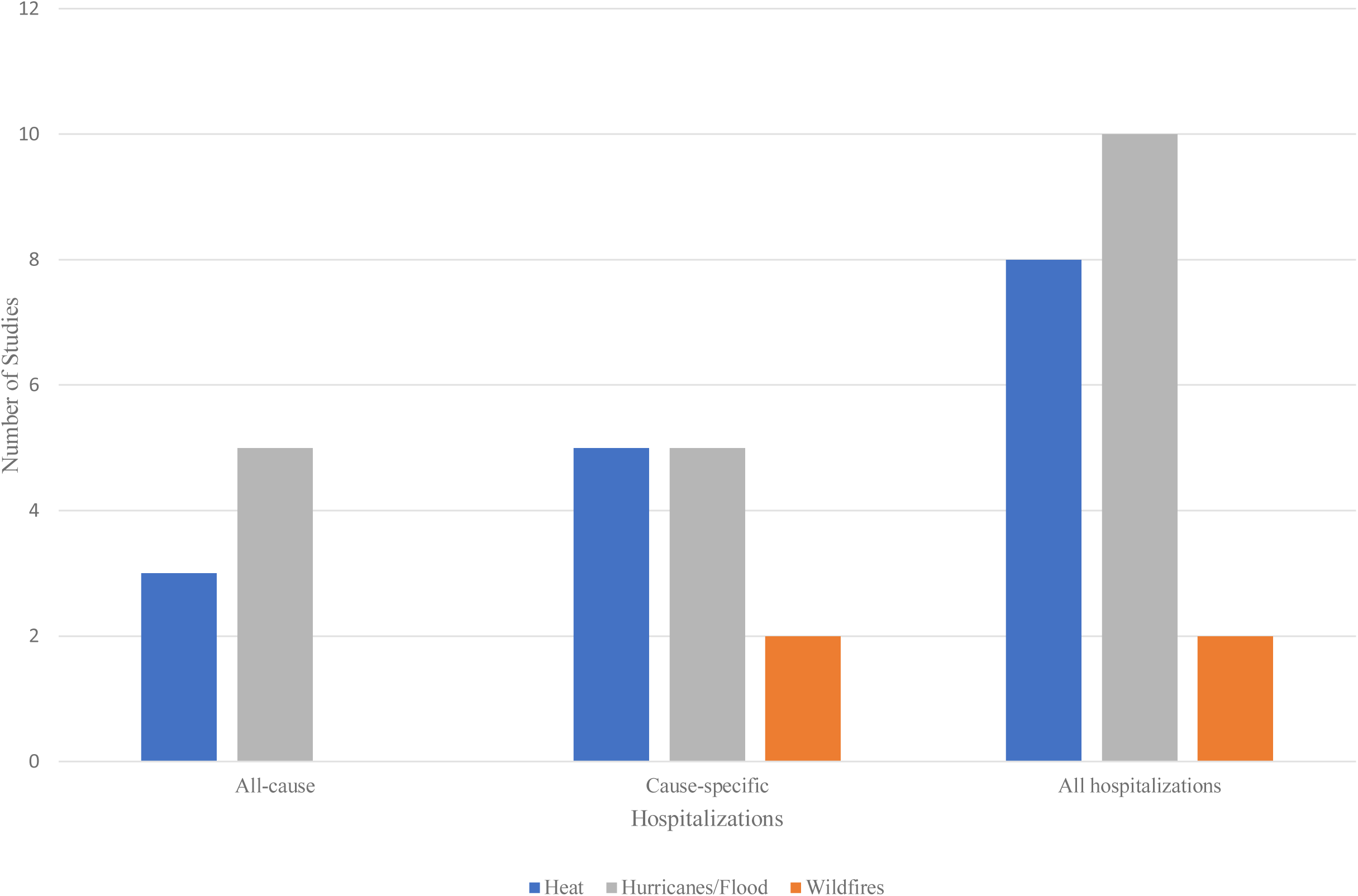
Hospitalizations by Climate-related Events. Footnote: illustrates the number of studies reporting hospitalizations associated with climate-related events, categorized by all-cause hospitalizations, cause-specific hospitalizations, and total hospitalizations. Hurricanes and floods were the most frequently studied events in relation to hospital admissions, followed closely by heat-related hospitalizations. Cause-specific hospitalizations were evenly reported for heat and hurricanes/floods, while wildfires were less frequently associated with hospitalizations in the reviewed studies.

#### Cause-Specific Hospitalizations

Of the studies reporting on hospitalizations **shown in** Figure 9, 12 examined cause-specific hospitalizations.^40,45,48,49,57,68,71,72,74,83,89,92^ Seven studies focused on hospitalizations due to respiratory causes,^45,68,71,72,83,89,92^ while four examined heatstroke/other heat-related conditions^40,48,49,57^. Most (n=4) of the studies examining respiratory causes of hospitalizations, such as those related to asthma, did so in the context of hurricanes/floods. ^72,83,89,92^ All studies examining heatstroke or other heat-related conditions were conducted in the context of extreme heat-related climate events. ^40,48,49,57^

**Figure 9.**
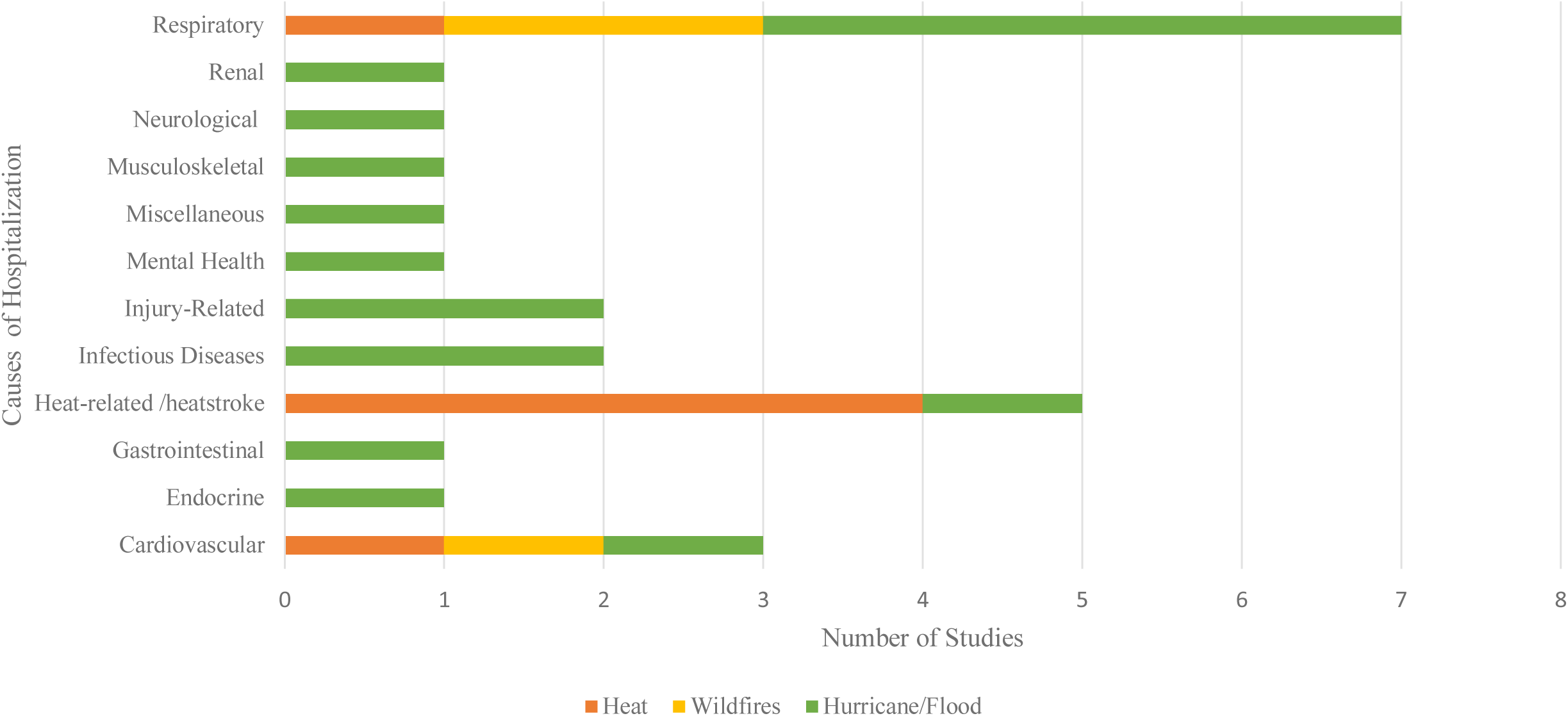
Cause Specific Hospitalizations. Footnote: presents the distribution of cause-specific hospitalizations associated with climate-related events, categorized by organ-specific health impacts. Respiratory conditions were the most frequently studied cause of hospitalizations, particularly in connection with hurricanes and floods, followed by wildfires, likely due to smoke exposure and poor air quality. Heat-related illnesses, including heatstroke, were most frequently reported in studies on extreme heat events, highlighting the direct health risks of rising temperatures. Cardiovascular conditions were also linked to both heat and hurricane/flood events, suggesting that extreme weather exacerbates underlying heart conditions. Other notable hospitalization causes included infectious diseases, mental health disorders, and injury-related cases, primarily associated with hurricanes and floods.

#### Health Care Utilization: Outpatient Services

Two studies reported on the effects of heat and wildfires on outpatient health care services.^44,70^ The first study examined the impact of heat on clinic attendance rates for those living with the human immunodeficiency virus (HIV) over a 383-day study period.^44^ The second study investigated the association between wildfire smoke exposure and asthma-specific medical care utilization in Oregon during the 2013 wildfire season (i.e., May 1, 2013, to September 30, 2013). This study used four metrics to characterize health care utilization: outpatient services, office visits, ED visits, and asthma rescue inhaler refill history. The first study found that HIV clinic attendance rates declined on days when the heat index was extreme compared to days with lower heat indices.^44^ The second study found that exposure to wildfire smoke in 2013 was associated with an increased risk of asthma diagnosis at EDs during the 22-week wildfire season (odds ratio [OR]: 1.089, 95% confidence interval [CI]: 1.043-1.136), office visits (OR: 1.050, 95% CI: 1.038-1.063), outpatient visits (OR: 1.065, 95% CI: 1.029-1.103) and asthma rescue inhaler medication fills (OR: 1.077, 95% CI: 1.065-1.088).^70^

#### Health Care Utilization: Undefined Site of Services

Our review identified one study that examined health care utilization related to diabetes screening following Hurricane Katrina. The study, using data from the Medicare Master Beneficiary Summary File and Personal Summary File from 2002-2004 and 2006-2008, analyzed claims for all Medicare beneficiaries enrolled in traditional Medicare. Although the specific site of care was not determined, the findings revealed a decrease in diabetes screening in the three years following Hurricane Katrina. This decline was most pronounced among older beneficiaries.^90^

#### Cost Implications of Climate-Related Events

Our review identified two studies^88,92^ that examined the impact of climate-related events on health care costs. The first study analyzed medical claims for 746,951 patients aged 65 and over treated at ED and inpatient facilities in New Jersey during November 2012, after Hurricane Sandy; claims data were compared to pre-Sandy baseline data in 2010 and 2011, and claims in 2013, 1 year following Hurricane Sandy. The study found an average increase of 1,700 cases in November 2012 for the top 15 ED conditions, with daily increases of 57 cases. This surge, primarily involving trauma cases, led to an approximate $7.3 million increase in health care costs for November 2012, with trauma care alone accounting for around $4.0 million.^92^

The subsequent study investigated the utilization of behavioral health services and associated costs before and after the Baton Rouge, Louisiana, flood in August 2016. Analyzing claims data from 163,057 Medicaid beneficiaries, study findings revealed that visits for substance use disorders (33.6% for males) and depression (30% for females) increased by 66% and 44%, respectively, after the flood. This led to a 4% increase in claims and a 14 % rise in costs over the 10 months post-flood.^88^ These costs for the Medicaid program to deliver services in the 10 months following the flood were estimated at $21,074,914, with an average per-visit cost of $107. Prior to the flood, the total cost for visits paid by Medicaid was $7, 260 059, with an average visit costing $95.

### Understanding the Impact of Climate-Related Events on Health Care Utilization in Specific Disease Populations

Six studies examined the effect of climate-related events on specific disease populations.^44,48,52,56,90,91^ These included diabetes (n= 2),^90,91^ end-stage renal disease (ESRD) (n= 1),^56^ rheumatic conditions (n= 1),^52^ persons living with HIV (n= 1),^44^ and those with unspecified chronic conditions.^48^ The first of the two studies focused on persons living with diabetes and examined whether older adults with diabetes who were also affected by Hurricanes Katrina (2005) and Rita (2005) had a greater number of ED visits and days hospitalized in the three years following Hurricanes Katrina (2005) and Rita (2005). Compared to a control group, those affected by the storms had an additional 380,907 days hospitalized and 21,583 ED visits.^91^ The second study examined health care utilization based on a predominantly older adult population with diabetes receiving a screen related to their disease; it found that in the 3 years following Hurricane Katrina (2005-2008), health care utilization was lower for those in the groups treated for their diabetes than in control groups, with the gap especially more pronounced for older adults.^90^

The single study of 7445 persons with ESRD, found that exposure to extreme heat events (defined by the study as calendar day- and location-specific 95th-percentile maximum temperature thresholds calculated using daily meteorological data from 1960 to 1989), was associated with an increased risk of same-day hospital admission (rate ratio [RR], 1.27; 95% CI, 1.13-1.43) and same-day mortality (RR, 1.31; 95% CI, 1.01-1.70) for patients with ESRD. In another study, among 14,401 persons with rheumatic conditions identified from the Mass General Brigham Research Patient Data registry between 2018-2021, individuals with rheumatic conditions living in areas with high versus low social and heat vulnerability had significantly greater odds of recurrent hospitalizations. ^56^

A study conducted at an HIV clinic in Miami, Florida, examined whether extreme heat days affected attendance rates. Analyzing 26,444 scheduled visits from 2017 to 2019, the authors found a significant increase in the relative risk of “no-show” status, a critical metric due to its impact on treatment adherence. Treatment adherence is crucial for controlling HIV, reducing complications, and preventing mortality. The study observed a 14% increase in no-show rates on days with an extreme heat index compared to days with lower heat indices, a 13% increase on days with extreme precipitation compared to days with no extreme rainfall, and a 10% increase on days with any extreme weather overall.^44^

Finally, one study examined the associations of heat-sensitizing medications (defined as medications used for chronic conditions, such as diuretics, and antipsychotics that disrupt thermoregulation or fluid/electrolyte balance, that make patients more sensitive to heat) and heatwaves with heat-related hospitalization among 9,721 older Medicare beneficiaries with at least one chronic condition.^48^

In this study, 42.1% of patients experienced a heatwave, and heatwaves were associated with an increase in heat-related hospitalizations ranging from 21% (95% CI: 7% to 38%) to 33% (95% CI: 14% to 55%) across medication classes. Several drug classes were associated with a moderately elevated risk of heat-related hospitalization in the absence of heatwaves, with rate ratios ranging from 1.16 (95% CI: 1.00 to 1.35) to 1.37 (95% CI: 1.14 to 1.66).^48^

#### Understanding The Duration of The Impact of Extreme Weather Events on Health Care Utilization

Few studies in our review report on the long-term consequences of these events regarding health care utilization. Among those that did, four focused on heat,^41,43,57,59^ one on wildfires,^61^ and six on hurricanes/floods.^73,76,81,89–91^

### Implications of Extreme Weather Events on Health Equity

Despite the heterogeneity across studies in measuring the health equity implications of climate-related events, some common themes emerged. Studies focusing on extreme heat frequently highlight age and racial-ethnic differences in health care utilization. Black individuals and those over 65 years old face heightened risks of heat-related health issues, leading to increased health care utilization. Historical inequities, such as racial segregation and redlining, have resulted in Black and other persons of color residing in U.S. regions, particularly in the Southern U.S., where they are more likely to encounter extreme heat.^94^ Additionally, Black individuals and other persons of color are more likely to work in outdoor jobs, lack access to air conditioning due to affordability concerns, and suffer from chronic conditions like asthma, which are exacerbated by extreme heat and other climate-related events.^94^

Older adults are also more vulnerable to heat-related and other climate-related events due to more complex medical comorbidities, higher rates of disability, and physiological changes.

Social factors such as housing insecurity and poor housing quality, including limited access to cooling systems, further increase the susceptibility of older adults to the effects of heat and other climate-related events.^95^

In the current review, regional disparities were evident, with the South, Northeast, and Midwest experiencing greater burdens of heat-related ED visits. One study also demonstrated higher health care utilization among rural populations in Montana due to extreme heat.^45^

Two studies examined health care utilization according to community Social Vulnerability Index (SVI) scores as defined by the Centers for Disease Control and Prevention (CDC). In the first study, communities in San Antonio, Texas, with high SVI scores—indicating greater disadvantage—had higher rates of EMS incidents.^54^ Another Massachusetts-based study found that individuals living in the highest SVI areas had 1.84 times higher odds (95% confidence interval [CI] 1.43-2.36) of experiencing four or more hospitalizations compared to those in the lowest SVI areas.^52^

Studies focused on wildfires have less frequently addressed health equity implications. Although some studies indicated a higher burden of wildfire-related health issues and health care utilization among specific demographic groups, these findings were not consistently stratified by demographic factors.

Studies focused on hurricanes/floods showed varied rates of inpatient visits by race and sex, with significant increases in health care utilization among Medicare populations. Vulnerable groups, including the oldest (85+), those living in poverty, and those with multiple comorbidities, were also found to have the highest probability of hospitalization following an extreme weather event. After disasters, lower-income populations experienced more new or worsening health issues, leading to increased use of health care services.

## Discussion

This rapid evidence review of 54 studies published between 2018 and 2023 provides unequivocal evidence that extreme heat, wildfires, and hurricanes/floods drive a significant increase in health care utilization, including ED visits, hospitalizations, and outpatient care. Despite variations in methodologies, exposures, outcome measures, study populations, and study periods, the findings are consistent: climate-related disasters place an escalating burden on health care systems. While only two studies included cost data, the overwhelming rise in health care utilization signals a substantial economic burden on individuals, communities, and systems, reinforcing the urgent need for targeted research and policy interventions to mitigate these impacts.

Most studies have focused on extreme heat and hurricanes rather than wildfires, possibly because these events have a broader impact. Each year, millions of Americans experience extreme heat, hurricanes, or both. For example, in June 2024, approximately 100 million Americans were under hot weather advisories as temperatures soared above 90°F in many areas, particularly in the Northeast and South.^87^ Additionally, in 2024, five tropical cyclones (Beryl, Debby, Francine, Helene, and Milton) primarily affected residents in the Southern U.S., impacting millions of people.^5,90,96^ These events are easier to study because they are often accompanied by widespread heat alerts, advisories, and the significant structural damage and displacement that hurricanes cause, which facilitates the tracking of health and other outcomes.^93,94,97^ In contrast, wildfires, though causing extensive structural damage and burning millions of acres in the U.S. annually, directly affect fewer lives. This discrepancy may also be due to methodological challenges in data collection related to tracking the direct impact of wildfires. ^93,94,97^ Variations in such factors as air quality and individual susceptibility further complicate research on wildfires.^98,99^

Furthermore, we found that the impact of these extreme weather events is inequitable, disproportionately affecting Medicare beneficiaries, who are older and have more medical comorbidities, and other vulnerable groups, such as those in areas of high social vulnerability. This finding highlights the need for future research to focus on the disproportionate impact on vulnerable populations, guiding targeted interventions and resource allocation to mitigate these effects and minimize potential threats that could increase Medicare spending. Studies should also account for geographical variations in health care impact, as different regions may have different vulnerabilities and health care capacities, helping to tailor local health care preparedness and response plans.

In the literature, the most described relationship between health outcomes and extreme weather events has been with extreme heat.^37^ However, our review reveals no clear consensus on which heat indicator is best to measure extreme heat. Some studies used absolute temperature cutoffs, while others used different metrics. This lack of consensus affects the ability to accurately estimate heat-attributable ED visits, hospitalizations, and outpatient visits, potentially leading to a gross underestimation of the actual burden on health care systems and the economic impact.

We observed variations in health care utilization patterns by the type of weather event. For example, studies examining ED visits most often focused on wildfires, while those reporting on hospitalizations mainly were centered on hurricanes. Most of these ED visits and hospitalizations were due to cardiovascular or respiratory conditions. A potential explanation is that wildfires typically lead to immediate and acute health issues triggered by smoke, such as acute asthma exacerbations, bronchitis, or other respiratory distress, which could warrant a visit to an ED or Urgent Care Center. In contrast, hurricanes can cause more severe physical trauma and exacerbation of chronic conditions due to disruptions in medical care, power outages affecting medical devices, and other stress-related health issues, which may require more extended periods of observation in a health care setting. Study design may also influence these differences in utilization patterns.

### Why are the Findings of This Study on Climate-Related Extreme Weather Events and Health Care Utilization Important?

1. Supports a need for health system planning to manage increased visits after climate events.
2. Calls for public health interventions to improve air quality and ensure clean water access post-events.
3. Emphasizes resource allocation to strengthen high-risk health systems.
4. Guides researchers in designing studies focused on event-specific health outcomes.
5. Advocates for better methods to link exposures and health care data, improving estimates and study comparability.
6. Highlights the need to expand literature reviews on health beyond heat-related events.
7. Calls for a consensus on a standardized heat metric.
8. Identifies common physical and mental stressors for specific events to enhance readiness.
9. Encourages research on mental health, chronic diseases, and outpatient utilization.
10. Emphasizes the need to quantify health care utilization and perform cost analyses— including infrastructure repair costs—to evaluate the cost-effectiveness of interventions for health care systems and governments.

### What are the Gaps Identified by this Literature Review?

11. Limited focus on outpatient service utilization, despite these settings providing 85-90% of health care services.1
12. Lack of data on outpatient care limits understanding of health care utilization and costs during climate events.
13. Insufficient research on specific disease populations, such as diabetes, ESRD, HIV, and cancer, which require ongoing care and are vulnerable to disruptions.
14. Need for studies examining mental health outcomes related to climate events.
15. Absence of consistent tracking of health care utilization and costs to evaluate the system’s resilience and preparedness for increased demand.
16. Lack of analysis on the economic impacts of climate events, including short- and long-term costs and their influence on insurance premiums.
17. Limited data to guide interventions for maintaining chronic disease management, preventive care, and mental health services during extreme weather events.

### Study Strengths and Limitations

This rapid review has several strengths. First, it contributes to the limited existing data that quantifies the health care utilization and economic burden associated with extreme weather events. Integrating findings across multiple studies provides a more comprehensive picture of the overall impact compared to previous studies that often focused on specific weather events or geographical regions. Second, the review underscores the importance of addressing vulnerable populations who face greater risks from extreme weather events. This focus is critical, as the health equity implications of such events have become central to public health efforts aimed at reducing health disparities. Third, the review identifies the need for future research, especially given the limited number of studies reporting on long-term health care utilization following a climate-related disaster. Last, the review underscores the necessity for more cost analyses, as only two studies reported on this aspect, highlighting the economic impact of climate-related events and the associated demand for health care services.

Despite these strengths, our study has limitations. We limited our review to three main extreme weather events, excluding less common events such as tornadoes, earthquakes, and droughts. Additionally, we restricted our synthesis to U.S.-focused studies to account for geographical differences in event frequency and variations in health care utilization and cost estimation methods, which differ based on health insurance and reimbursement structures in other countries. Furthermore, we only included studies published in the past five years to capture the most recent data. As a rapid evidence review, we did not conduct an exhaustive search of the literature, meaning some relevant articles published within the date range and available in databases other than PubMed may have been omitted. Despite these limitations, we believe the findings, particularly the emphasis on providing a call to action and identifying outstanding research questions, represent a valuable contribution to the field.

### Policy Strategies for Mitigating the Health Impacts of Climate Change

The climate crisis is no longer a future threat—it is a present emergency straining health care systems, worsening outcomes, and deepening inequities. These impacts are accelerating, indifferent to political leadership or partisan boundaries. While evidence linking extreme weather events to public health continues to grow, there is still an urgent need for bold policies that both mitigate climate change and strengthen health systems’ preparedness. Between 2021 and 2024, the Biden administration made significant strides in advancing climate research and policies, including launching the Climate Change and Health Initiative with $40 million in annual funding in 2023 and 2024, and passing the Inflation Reduction Act (IRA) (2022), which supported clean energy, improved health infrastructure, and boosted resilience in vulnerable communities.^100^

Several federal initiatives under the Biden administration also addressed the intersection of climate and health. A 2021 Executive Order integrated climate into U.S. foreign policy and national security, directing agencies to cut emissions, boost resilience, and promote environmental justice through clean energy and job creation.^101^ That same year, the Department of Health and Human Services launched the Office of Climate Change and Health Equity (OCCHE), which has since led initiatives like the 2024 IRA Catalytic Program to help safety net providers build climate resilience.^102^ In 2024 alone, the CDC introduced a national Heat and Health Index Tracker, ^93^ Department of Labor’s Occupational Safety and Health Administration (OSHA) proposed the first federal workplace heat safety standard,^103^ and the Securities and Exchange Commission (SEC) finalized a rule requiring disclosure of climate-related financial risks by investors. ^104^

Under Ranking Member Richard Neal, the House Ways and Means Committee has taken key steps to address the climate crisis’s impact on health care. This includes a 2022 congressional hearing and multi-part reports outlining how extreme weather affects health care systems, how organizations are reducing their carbon footprints, and how groups like group purchasing organizations (GPOs) are responding.^19,20,105^ These efforts are vital as millions of Americans face the growing toll of climate-driven events each year.

In stark contrast, the current Trump administration has begun rapidly dismantling key federal climate initiatives. Executive actions have led to the U.S. withdrawal from the Paris Climate Accord—a cornerstone of global climate cooperation—and, on January 20, 2025, rescinded Executive Order 14008, which had established critical offices and councils to coordinate domestic and international climate action.^106^

Following the rescission of Executive Order 14008 in January 2025, the Trump administration has intensified efforts to dismantle the federal climate agenda. In early 2025, over 1,000 staff were laid off at the National Oceanic and Atmospheric Administration (NOAA) and hundreds more at the Federal Emergency Management Agency (FEMA), alongside escalating threats to eliminate FEMA entirely — moves that would severely undermine the nation’s ability to respond to climate-driven disasters.^107,108^ At FEMA, more than $100 billion in previously awarded grant funds and disaster assistance remains frozen.^108^ Simultaneously, research grant funding was halted across multiple agencies, including the National Institutes of Health (NIH), and deep cuts were made within the Department of Health and Human Services (HHS). These actions culminated in the closure of OCCHE, effectively halting critical IRA initiatives designed to address the health impacts of climate change.^102^

In addition to the workforce impacts, the Trump administration has also proposed cutting NOAA’s competitive climate research grants program. A move that would end support for collecting regional climate data and result in the loss of nearly $70 million a year in funding to academic scientists conducting climate-related research.^109^ Such a move would be devastating not only to the health care sector but across multiple industries, such as the agriculture sector, which rely on this data. The Trump administration also proposes to terminate NOAA’s National Oceanographic Partnership Program and college and aquaculture sea grant programs, which would undermine and essentially halt critical research in these areas.^109^

In March 2025, the NIH announced it would no longer fund research on the health impacts of climate change.^110^ Princeton University alone lost over $4 million in federal grants, including Department of Commerce funding for a five-year study on how global warming affects Earth’s water availability.^111^ Funding and staffing for the National Climate Assessment, a congressionally mandated report—responsible for tracking climate’s impacts on health, agriculture, and the economy—have also been slashed.^112^ States’ climate actions are now being actively targeted by the Trump administration following the issuance of the *Protecting American Energy from State Overreach* Executive Order on April 8, 2025.^113,114^

In April 2025, the Trump administration canceled $1 billion in funding to Cornell University and $790 million to Northwestern University—both leaders in climate and health research—citing alleged civil rights violations.^115^ Nearly 60 additional universities now face potential funding cuts under similar claims.^115,116^

Climate-driven disasters—heatwaves, wildfires, floods, hurricanes—are becoming increasingly frequent, severe, deadly, and costly. In the face of political backsliding, the research community has both a moral and scientific responsibility to confront these escalating threats. Rolling back climate policy does not halt the crisis—it only leaves communities more vulnerable and less prepared when disaster strikes. In the absence of sustained federal leadership, this moment presents a critical opportunity to forge new leadership through cross-sector collaboration.

Academic institutions, health systems, and the private sector must come together to drive climate and health research, track and report health impacts, and develop targeted interventions that strengthen resilience. The path forward demands urgent, coordinated action to ensure all communities—especially the most vulnerable—are equipped to withstand the growing health consequences of the climate crisis.

### Future Research Questions

- How do health care utilization and costs change over time following climate-related events?
- What is the impact of climate-related events on specific groups or disease populations?
- How can health care systems improve their resilience to climate-related events?
- How can health care systems reduce carbon emissions and shift toward greener practices to mitigate their contribution to climate-related events that harm public health?

**1. How do health care utilization and costs change over time following climate-related events?**

Future research should focus on longitudinal studies to track changes in health care utilization and costs over extended periods following climate-related events. This will help identify long-term trends and the sustained impacts on health care systems, provide insights into the effectiveness of current mitigation strategies, and highlight areas for improvement.

**2. What is the impact of climate-related events on specific groups or disease populations?**

There is a need for more research examining the differential impacts of climate-related events on specific groups, such as older adults, children, individuals with chronic health conditions, those with disabilities, or individuals from other marginalized backgrounds. Studies should explore how these populations are affected differently and identify targeted interventions to support their unique needs. Additionally, having a disease focus would allow for the examination of the impact of extreme heat on heat-sensitive drugs.

**3. How can health care systems improve their resilience to climate-related events?**

It is crucial to investigate the effectiveness of various strategies and interventions aimed at enhancing the health care system’s resilience. Research should focus on evaluating existing preparedness and response plans, identifying best practices, and developing new approaches to ensure that health care services can continue to operate effectively during and after climate-related events.

**4. How can health care systems reduce carbon emissions and shift toward greener practices to mitigate their contribution to climate-related events that harm public health?**

Addressing this question allows for the examination of the crucial role of the health care sector in combating climate change by identifying strategies for reducing carbon emissions, implementing sustainable practices, and leveraging the sector’s influence to drive broader environmental improvements. Decarbonizing health care not only helps mitigate climate impacts but also sets a leading example for other industries to follow in reducing environmental harm.

**5. Which federal interventions should be further evaluated and better leveraged to address climate-related health challenges? Specifically, given the disproportionate impact of climate events on older adults and the high enrollment in Medicare Advantage plans, how do Medicare Advantage supplemental benefits—such as air conditioning and structural home modifications for vulnerable populations—align with the geographic distribution of extreme weather events driven by climate change?**

Addressing this question is crucial because, while Medicare Advantage supplemental benefits aim to address social determinants of health, they often fail to reach the most vulnerable populations who need them the most. Given that these groups are also more susceptible to climate-related events, it is crucial to understand if benefits, such as air conditioning, home modifications, and other forms of living support, are available and accessible to those with the greatest needs. Gaining this understanding can help shape policies that ensure a more equitable distribution of resources, particularly as climate-driven weather events become more frequent and severe.

Additionally, other federal health care programs should be examined through a similar lens to determine how best to allocate funding to improve health outcomes and mitigate climate-related impacts. For example, the Low-Income Home Energy Assistance Program (LIHEAP), administered by the HHS, provides financial assistance to low-income households for energy costs, including cooling and heating.^117^ Evaluating how LIHEAP resources are distributed in climate-vulnerable regions could offer insights into optimizing federal spending to enhance health resilience in the face of climate change. By exploring these research questions, the evidence base can be developed to improve understanding of the complex relationship between climate change and health care, ultimately contributing to more effective and sustainable solutions to protect public health in an evolving climate.

### Future Challenges for Researchers Engaged in Climate-Related Research

As the climate crisis intensifies, researchers and practitioners working at the intersection of climate and health have a critical role to play in establishing new leadership. However, doing so requires confronting several persistent methodological and data-related challenges. Chief among these is the difficulty of measuring causal relationships between climate events and health outcomes, particularly over time. Longitudinal analyses are essential to understanding not only acute impacts but also how climate-related exposures influence health care utilization, costs, and outcomes across diverse populations. Data availability and quality remain significant obstacles. For example, while Medicare data from the Centers for Medicare & Medicaid Services (CMS) offer valuable insight, access to information on Medicare Advantage enrollees—who now comprise the majority of Medicare beneficiaries^118^—is limited. These data are managed by private insurers, are often proprietary and non-standardized, and are far less accessible than traditional Medicare fee-for-service claims. This lack of transparency presents a significant barrier to understanding the full burden of climate-related health impacts among older adults and individuals with disabilities, populations known to be especially vulnerable.

Capturing the cumulative and compounding effects of climate events is another challenge. Individuals may experience multiple overlapping exposures—such as extreme heat followed by hurricanes or wildfires—which complicates attribution and analysis. In many cases, health outcomes may not be immediately apparent or easily linked to a single event. Moreover, those who evacuate or avoid the direct impact of a disaster often face significant health consequences, which must also be accounted for in research.

Addressing these complex challenges will require innovative methodologies, improved data sharing, and collaborative frameworks across sectors. Only then can the research community generate the robust evidence needed to inform policy and protect public health in a rapidly changing climate.

## Conclusion

The evidence is clear: extreme weather events are linked to increased health care utilization and rising costs. Yet critical gaps remain—particularly around the long-term impacts, differential effects on vulnerable populations and disease groups, and the actual financial burden on the health care system. Older Americans, many of whom are Medicare beneficiaries, are especially at risk, underscoring the urgency of targeted, data-informed interventions.

In fiscal year 2024 alone, the U.S. government spent approximately **$1.9 trillion** on health care programs and services.^119^ Mitigating the health impacts of the climate crisis is not only a moral imperative—it is a fiscal one. Strategic action can improve outcomes while reducing long-term costs, especially among those most vulnerable to climate-related harms.

However, this progress is now under threat. The recent wave of political rollbacks—gutting federal climate initiatives, cutting critical research funding, and dismantling agencies tasked with climate preparedness—places the nation at greater risk.

This moment demands more than acknowledgment—it requires bold, coordinated action. A comprehensive policy framework must prioritize cross-sector collaboration, protect funding for climate-health research, strengthen data infrastructure, and ensure equitable access to climate-resilient health care systems. Researchers, health professionals, and community leaders must collectively lead this charge—translating science into solutions, and advocacy into impact—to safeguard public health in the face of a rapidly accelerating climate crisis.

## Funding Statement

The author received no funding for this study.

## Disclosures

The author declares no relevant conflicts of interest to disclose.

## Data availability

Data sharing is not applicable to this article as no datasets were generated or analyzed during the current study.

## Ethics approval and consent to participate

Not applicable

## Consent for publication

Not applicable

## Availability of data and materials

This rapid evidence review was conducted using data aggregated across already published studies

## Conflicts of Interest

The author declares there are no conflicts of interest

## Funding

The author received no financial support for the research, authorship, and/or publication of this article.

## Contributions

**Conceptualization:** Shakira J. Grant, Rachel Dolin

**Data curation:** Shakira J. Grant, Ari Panzer

**Formal analysis**: Shakira J. Grant, Ari Panzer

**Funding acquisition:** Not applicable

**Investigation:** Shakira J. Grant, Ari Panzer, Rachel Dolin

**Methodology:** Shakira J. Grant, Ari Panzer, Rachel Dolin

**Project administration**: Shakira J. Grant

**Resources**: Shakira J. Grant

**Software:** Shakira J. Grant

**Supervision:** Shakira J. Grant

**Validation:** Shakira J. Grant, Ari Panzer

**Visualization:** Shakira J. Grant, Ari Panzer

**Writing** – original draft: Shakira J. Grant

**Writing** – review & editing: Shakira J. Grant, Ari Panzer, Rachel Dolin, Amy Hall, Sarah Levin

## Acknowledgments

This work was conducted while the primary author, Shakira J. Grant served as a Health Policy Advisor/Fellow on the Democratic Staff of the U.S. House Committee on Ways and Means. The author gratefully acknowledges the contributions and support of committee staff members, Amy Hall, Sarah Levin, Rachel Dolin, and Ari Panzer. Additional support was provided by the Congressional Research Service, which conducted the initial literature search and compiled a list of relevant studies. We also thank Ashley Rodgers, former Fellow on the Ways and Means Health Subcommittee, for her assistance with data abstraction. Finally, we acknowledge the contributions of Ayesha Khan, Sierra Kaplan, and Grace Wang, summer interns on the committee for their work in assembling project tables.

## Notes

### Competing Interest Statement

The authors have declared no competing interest.

### Funding Statement

The study did not receive any funding.

### Author Declarations

The study utilized data from studies previously published in scientific journals.

